# A Mobile App for Prolonged Grief among Bereaved Parents: Study Protocol for a Randomized Controlled Trial

**DOI:** 10.1101/2021.04.23.21256003

**Authors:** Rakel Eklund, Maarten C. Eisma, Paul A. Boelen, Filip Arnberg, Josefin Sveen

**Author notes:** Corresponding author: Rakel Eklund, Uppsala University, Department of Neuroscience, SE-751 85 Uppsala, Sweden.

## Abstract

**Introduction:** Bereaved parents have elevated risk to develop mental health problems, yet, few studies have evaluated the effect of psychosocial interventions developed for bereaved parents. Cognitive behavioral therapy (CBT), both face-to-face or digitally delivered, has shown to be an effective intervention for prolonged grief symptoms. Self-help mobile apps offer various advantages and studies show improved mental health after app interventions. No app has yet been evaluated targeting prolonged grief in bereaved parents. Therefore, the aim of this planned study is to develop and examine the effectiveness of a CBT-based mobile app, called *My Grief*, in reducing symptoms of prolonged grief, as well as other psychological symptoms, in bereaved parents. Another aim is to assess users’ experiences and adverse events of *My Grief*.

**Methods and analysis:** We will conduct a two-armed randomized waitlist-controlled trial. Parents living in Sweden, who lost a child to cancer between one and ten years ago, with elevated symptoms of prolonged grief, will be recruited to participate in the trial. The content of *My Grief* covers four main domains (Learn; Self-monitoring; Exercises; Get support) and builds on principles of CBT and the proven-effective PTSD Coach app. Participants in the intervention group will fill out online questionnaires at baseline and at 3-, 6- and 12-months follow-ups, and the waitlist-controls at baseline and at 3 months. The primary outcome will be prolonged grief symptoms at the 3 months follow-up. Secondary outcomes are posttraumatic stress and depression symptoms, quality of life, and cognitive behavioral variables (i.e., avoidance, rumination, negative cognitions).

**Ethics and dissemination:** Ethical approval has been received from the Swedish Ethical Review Authority (project no. 2021-00770). If the app is shown to be effective, the app will be made publicly accessible on app stores, so that it can benefit other bereaved parents.

**Trial registration:** Clinicaltrials.gov, identifier: NCT04552717.

**STRENGTHS AND LIMITATIONS:**

- This is the first study to examine whether access of a self-help app can have beneficial effect on bereaved parent’s mental health, quality of life
- This study will examine the effect of the app on cognitive-behavioral processes proposed to underlie the development of prolonged grief.
- Generalizability of findings from this study may be limited as parents who want to participate in such study may experience fewer barriers to talk about the loss and seek help for their grief.
- This study includes parents who have lost a child to cancer, hence the findings may not be generalizable to other causes.
- This study uses self-report questionnaires, hence we do not establish formal diagnosis of prolonged grief disorder.

## INTRODUCTION

The death of one’s child can be a traumatic life experience and bereaved parents are at increased risk of developing both mental and physical health problems (1–6).

While most bereaved individuals adjust to the loss of a loved one without professional intervention, a significant minority experience a persistent, severe and disabling grief reaction, called prolonged grief. Prolonged Grief Disorder (PGD), a diagnosis characterized by such severe grief responses, was recently included in the International Classification of Disorders eleventh edition (7). A similar version of PGD is scheduled to be included in the text revision of 5^th^ edition of the Diagnostic and Statistical Manual of Mental Disorders (DSM-5-TR for a commentary see Boelen et al., 2020 (8)). Across both disorders, core symptoms are persistent and pervasive yearning for the deceased, persistent and pervasive cognitive preoccupation with the deceased, as well as accessory symptoms presumed indicative of intense emotional pain, such as difficulty accepting the loss, a feeling of that one has lost a part of one’s self, and difficulty in engaging in social activities. Such severe grief reactions often co-occur with, yet are distinct from, disorders such as depression and posttraumatic stress (9), and relate to suicidal ideation (10), as well as reductions of quality of life (11).

Bereaved parents are at high risk to develop prolonged grief (12–14). For example, 16% of cancer bereaved parents may have reactions indicative of PGD (15). Yet, there is a lack of studies evaluating the effect of psychosocial interventions for bereaved parents (16,17). Thus, the development and evaluation of accessible interventions to prevent and reduce negative mental health consequences after child loss appears clinically important.

A growing body of evidence shows that cognitive behavioral therapy (CBT) is effective in treating prolonged grief symptoms when delivered face-to face (18–21), or via internet (22–24). Briefly, the cognitive-behavioral model of prolonged grief proposes that three core processes explain the occurrence and persistence of grief symptoms: insufficient integration of the loss with existing autobiographical knowledge; negative global beliefs about oneself, the world and the future and catastrophic misinterpretations of grief symptoms; and anxious avoidance (i.e., cognitive and overt avoidance of reminders of the loss) and depressive avoidance (i.e., avoidance of social, occupational and recreational activities, and behavioral withdrawal) (25). Accordingly, CBT treatments of prolonged grief typically includes creating a coherent, meaningful autobiographical narrative about the loss; challenging negative beliefs and catastrophic misinterpretations through cognitive restructuring; gradually confronting persons in vitro or in vivo with avoided aspects of the loss (e.g., places, objects, memories) through exposure techniques; and/or helping people to set new life-goals and engage in new, meaningful activities (26). Increasingly, researchers are also focused on examining the efficacy of other, complementary therapeutic techniques, which may be helpful in reaching these treatment goals, such as mindfulness-based therapy and relaxation (27–29).

Mobile apps offer additional advantage over face-to-face and web-based interventions because of their availability, accessibility, the immediate support they can provide, and their anonymity (e.g., reducing barriers to seeking help for grief), low costs, and possible tailoring to the user (e.g., the user are in control over when and how to use the app) (30). Most individuals own a smartphone, regardless of socioeconomic background or living conditions, and many spend 2-5 hours a day using their smartphone (31). Moreover, people with mental health issues report daily use of smartphones to connect to other people and search for health-related information (32,33). Hence, mobile apps have a great potential to be used as self-help interventions for mental health problems. Indeed, various mobile apps for mental health have shown to be effective (34,35). Treatment trials have shown that mobile apps can decrease a variety of psychological symptoms, including posttraumatic stress (36,37) but also depression (38), anxiety (39), and substance use (e.g., tobacco, alcohol and drugs) (40).

Despite the availability of a large number of mobile apps in app stores that claim to improve mental health, empirical evidence is lacking for most mobile applications (30,34,35). To our knowledge, no evidence-based app to reduce grief for bereaved parents exists. Based on the existing knowledge, we believe an app targeting prolonged grief using elements of CBT could potentially be effective in improving mental health in bereaved parents.

The general goal of the present intervention study is to evaluate the effectiveness of an app for bereaved parents, which aims to facilitate the grieving process. The intervention includes access to a self-management mobile app, called *My Grief*, and is based on the cognitive-behavioral conceptualization framework of prolonged grief (18,25). The intervention also includes other evidence-based techniques from CBT such as elements of mindfulness and deep muscle relaxation practices, as these techniques have shown promising effects on loss-related mental health for bereaved individuals (27).

The main aim of this study is to examine the effectiveness of the app *My Grief* as compared to a waiting list comparison in reducing symptoms of prolonged grief in bereaved parents. The primary hypothesis is that the parents in the intervention group will report decreased levels of prolonged grief symptoms after having access to the app. A second aim is to examine the effect of the app on related mental health problems (posttraumatic stress symptoms, depression symptoms, quality of life) and cognitive behavioral variables putatively explaining the effect of CBT techniques (i.e., grief avoidance, grief rumination and negative grief cognitions). The secondary hypotheses are that the parents in the intervention group will report improved mental health (i.e., lower symptom levels of depression and posttraumatic stress, higher quality of life), and lower grief avoidance, grief rumination and negative grief-related cognitions. A third aim is to assess participants experience of using the app, e.g., perceived satisfaction helpfulness, and adverse events.

## METHODS AND ANALYSIS

### Design

We plan to conduct a two-armed parallel-group randomized waitlist-controlled trial comparing the effects of access to the *My Grief* for three months compared to waitlist. Randomization will take place after the pre-assessment is completed, using a random number generator (www.random.org) by a blinded independent researcher, using block randomization with an allocation ratio of 1:1. Participants will be informed of group allocation immediately after randomization (Figure 1).

**Figure 1.**
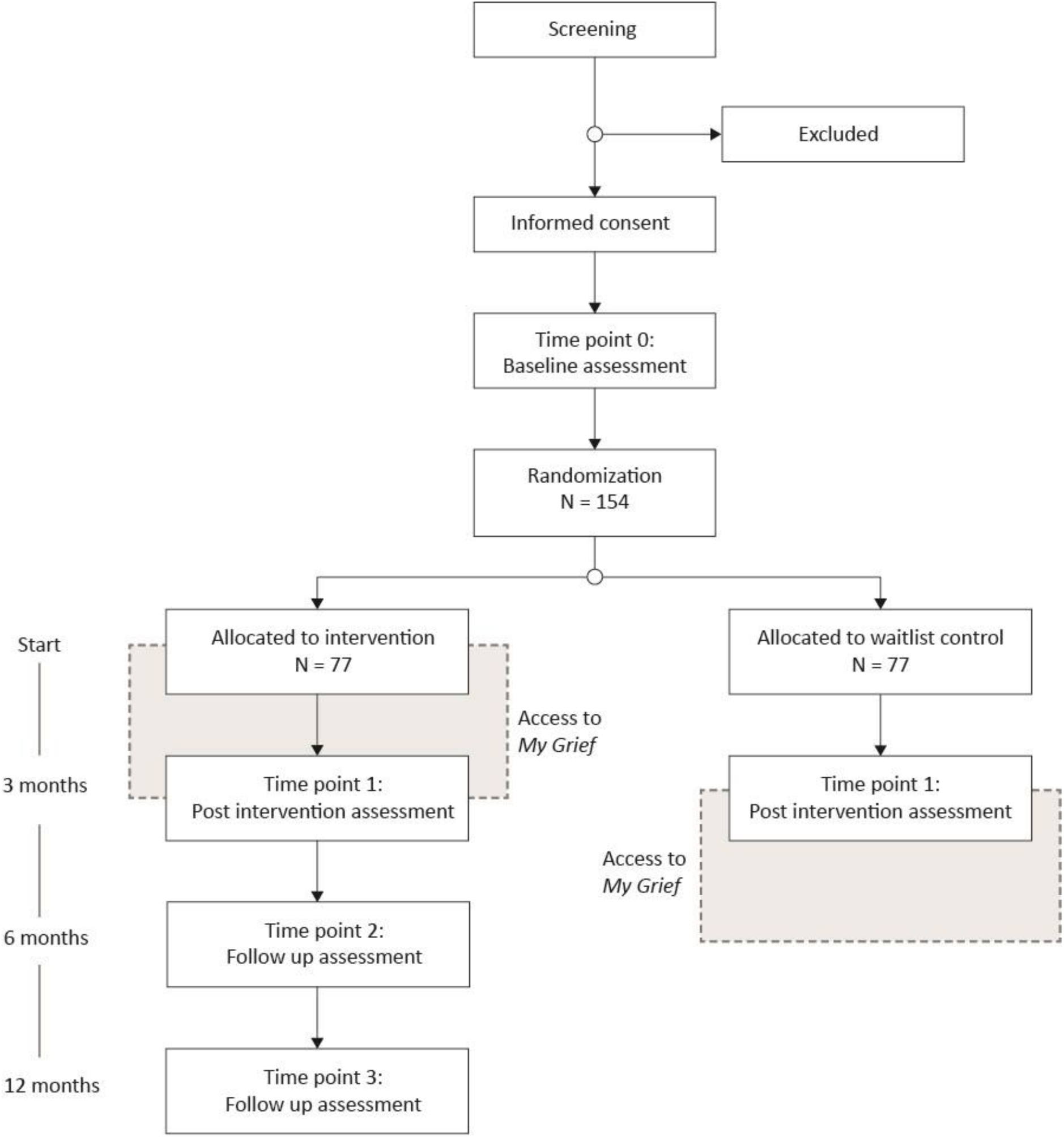
Design of the RCT

### Participants and procedures

Parents living in Sweden, who lost a child to cancer between 1 and 10 years ago, with elevated symptoms of prolonged grief, will be recruited to participate in the intervention. Potential participants are identified using the Swedish Childhood Cancer Registry, the Cause of Death Registry, and the Swedish Population Register at the Swedish Tax Agency. By linking the Cause of Death Registry with the Swedish Childhood Cancer Registry, children diagnosed with a malignancy and who died due to the malignancy 1 to 10 years previously will be identified. Next, the children’s parents are identified through the Swedish Population Register and will be sent a letter with an invitation to participate in the study. People who are interested in participation are directed to a website, www.minsorg.com, where they can access information about the study aims and procedures. On the web-site they can sign up to the study and complete an online screening questionnaire which automatically direct eligibly participants to an informed consent form and the pre-assessment online questionnaire, in REDcap (41). After filling in the pre-assessment they are randomized by an independent person to either the intervention or waiting list group. Participants in the intervention group will receive a phone call from a researcher one week after the intervention started to ensure that the app is working properly. Participants in the intervention group are allowed to use the *My Grief* app for three months. The control group will be informed that they are on waiting list and will be offered access to the *My Grief* app after three months (Figure 1). Recruitment is planned to start August 2021.

### Eligibility criteria

Inclusion criteria for participation are: 1) being a parent of a child who has died of cancer between 1 and 10 years previously, 2) having elevated symptom levels of prolonged grief, 3) understanding and speaking Swedish, and 4) having access to a smartphone. Exclusion criteria are self-reported ongoing suicidal thoughts or psychosis, assessed with single-items in the screening questionnaire.

### Sample size

A required sample size of 154 participants was calculated in G*Power version 3.1, based on the primary research question. We considered a moderate effect size to be clinically meaningful (*f* = 0.25), within a repeated measure ANOVA, a desired power of 0.80, *α* = 0.05, and an assumed strong association (*r* = 0.50) between the pre-assessment and post-assessment. The estimate includes an anticipated attrition rate of 16% (42).

### Intervention

*My Grief* is a self-help app for smartphone users, compatible with both iOS and Android. The app builds on principles of CBT and a PGD online-treatment protocol (43). It is based on a cognitive-behavioral conceptualization of prolonged grief and proposes that three “grief tasks” are critical in alleviating grief (18,25). These tasks are 1) facing the loss and the pain that goes with it, 2) keeping confidence in yourself, others, life and future, and 3) engaging in helpful activities that promote adjustment to the new situation. Interventions used to accomplish these tasks include psychoeducation and normalization of grief reactions, exposure to avoided aspects of the loss, and goal setting and behavioral activation. The structure of the app includes four main sections (Figure 2) and is based on PTSD Coach app with permission from the Veterans’ Affairs National Center for PTSD and Department of Defense’s DHA Connected Health (36,44).

**Figure 2.**
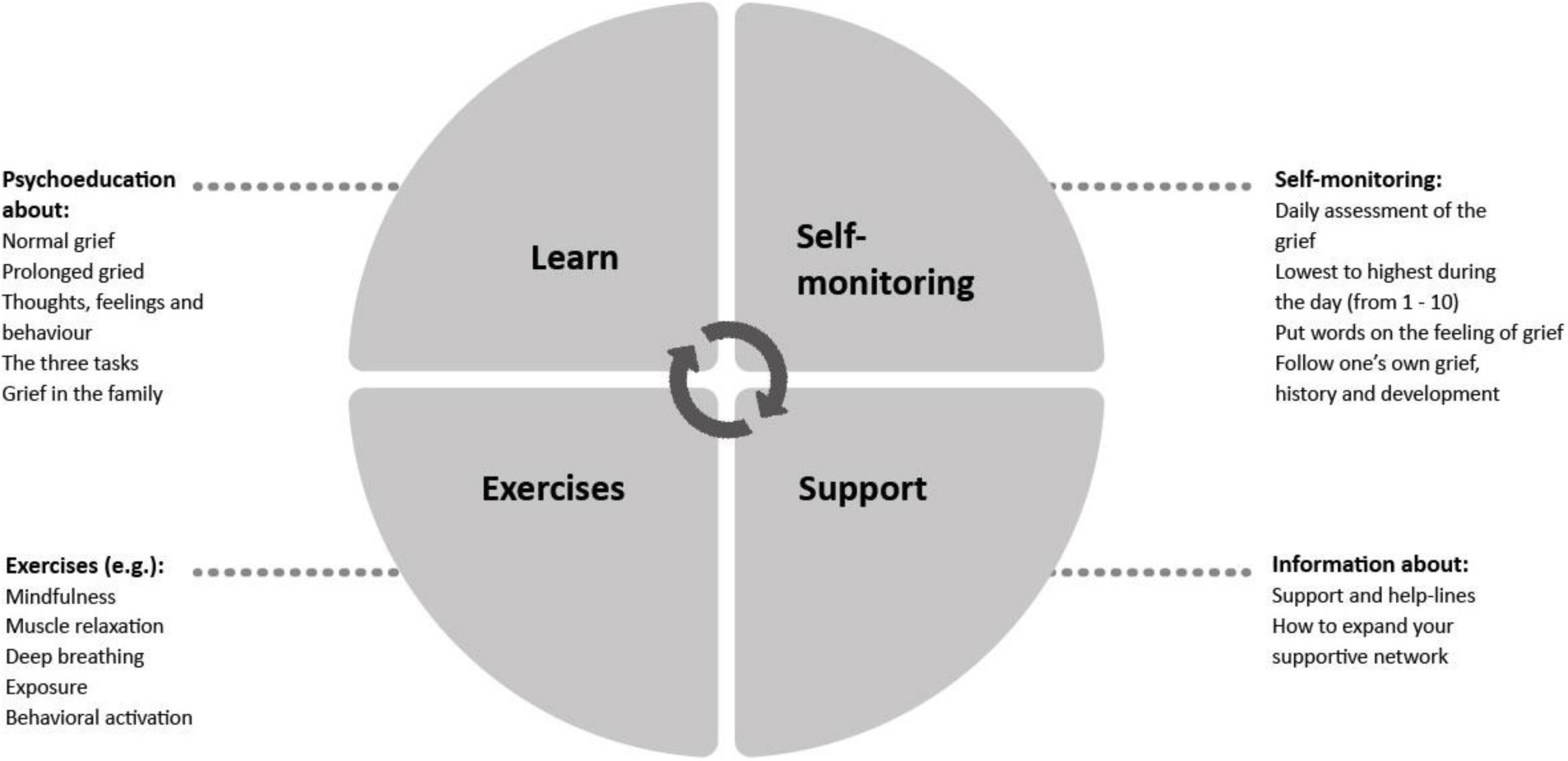
Structure of the *My Grief* app.

The four main sections in *My Grief* include:

1. *Learn*. This section provides psychoeducation about grief. It covers what is normal and prolonged grief; what commonly experienced thoughts, feelings and behaviors are after loss; the three grief tasks; and coping with grief within the family. The latter topic includes tips on how to understand different family members’ grieving processes, how to help each other, how to continue doing things together, how to support the remaining children in the family (i.e., siblings to the deceased child) in their grief process, and how to talk to children about death. In each part, there are links to corresponding exercises in section 3.
2. *Self-monitoring*. This section includes a rating scale (score 0-10) that users can employ to assess the daily intensity of their grief and the highest and lowest grief levels during the day. It also provides space to note on what happened at that time. The section gives users both visual and written feedback from themselves as the results are presented together with previous assessments. This enables users to monitor their grief intensity and symptoms over time, to recognize situations when grief levels are low or high, and to identify activities and internal and external stimuli associated with increases and decreases in grief; this helps them to learn more about one’s own grief and to improve adaptive ways of coping. User can use the rating scale as many times as they want, whenever they want. Participants are encouraged to fill in the scale daily and they can choose to receive a daily reminder to fill in the scale.
3. *Exercises*. This section provides the user with self-guided exercises. A majority of the exercises include recorded audio. For example, the user can choose to read or listen to instructions for a particular exercise. Some of the exercises were taken from the PTSD Coach Sweden (45), including mindfulness exercises (e.g., body scan, mindful breathing, learning to be aware of one’s senses), progressive muscle relaxation, deep breathing, and stress reduction and positive psychology exercises. Other exercises were derived from CBT (18,25). The writing exercise section includes tasks covering the three grief tasks in the CBT model. Three writing exercises, drawing from an internet-based exposure treatment (43), are related to task one. In these writing exercises, participants expose themselves to the story of loss and are encouraged to confront implications of the irreversibility of the separation from the deceased. For example, in the exposure to the story of the loss the participants are instructed to write down what happened around the time of the death, in three steps, in order to confront feelings, thoughts and situations which are difficult and emotionally painful. One exercise is related to grief task two and instructs participants to every day take a few minutes to think positive about the future and what they want for the future and to write this down in a diary. One exercise related to grief task three, is based on behavioral activation principles. It consists of encouragement to plan a moment every day to engage in an activity that potentially elicits some pleasure or joy.
4. *Get support*. This section provides the user with contact details to different support functions and help-lines (including phone numbers, chats, e-mail addresses, and websites) for both the parent and his/her family members. These support functions or help-lines include the Childhood Cancer Foundation, a suicide prevention organization, and different networks for parents who lost a child. It also includes psycho-education regarding how the user can create and expand one’s own support network throughout reaching out for help to family and friends. Participants can create a list of phone-numbers to family, friends and help-lines in this section as well.

### Data collection

Participants complete questionnaires online. Time points for data collected via online questionnaires at t0 (baseline), t1 (3 months), t2 (6 months, only intervention group) and t3 (12 months, only intervention group) (Table 1; Figure 1). If the participant has not completed the online questionnaire, two reminder emails will be sent out for each measurement time-point.

**Table 1.** Timepoints for measurements

|  | Before baseline | t0, baseline | t1, 3 months | t2, 6 months<sup>a</sup> | t3, 12 months<sup>a</sup> |
| --- | --- | --- | --- | --- | --- |
| Consent | X |  |  |  |  |
| Screening | X |  |  |  |  |
| Sociodemographic |  | X |  |  |  |
| Prolonged grief (PG-13) |  | X | X | X | X |
| PTSD (PCL-5) |  | X | X | X | X |
| Depression (PHQ-9) |  | X | X | X | X |
| Quality of life (BBQ) |  | X | X | X | X |
| Grief avoidance (DAAPGQ) |  | X | X | X | X |
| Grief rumination (UGRS) |  | X | X | X | X |
| Negative grief cognitions (GCQ-18) |  | X | X | X | X |
| Users' experiences<sup>a</sup> |  |  | X |  |  |

<sup>a</sup>Only intervention group

#### Sociodemographic and loss-related variables

A self-constructed questionnaire will be used to gather information about the participants’ demographic information, such as age, gender, education, employment, the child’s form of cancer and date of the child’s death. These data will be used to describe the sample.

#### Primary outcome

*Prolonged grief symptoms* will be measured by “Prolonged Grief Disorder-13” (PG-13). It consists of 13 items including 11 items assessing cognitive, behavioral and emotional symptoms during the past month, rated on a 5-point scale (not at all – several times a day/overwhelmingly). It also contains two items on duration and impairment (yes/no). PG-13 is scored by summarizing the 11 items regarding symptoms. The total score ranges from 11 to 55, with higher scores indicates more severe PGD (46). The instrument is validated in a Swedish sample with bereaved parents and indicates satisfactory psychometric properties (15).

#### Secondary outcomes

*Posttraumatic stress disorder symptoms* will be measured with the Posttraumatic Stress Disorder Checklist for DSM-5 (PCL-5) at baseline and follow up. It consists of 20 items describing symptoms of posttraumatic stress. Participants rate on a 5-point scale (not at all – extremely, 0-4) to what extent they experience symptoms. The total score ranges from 0-80, and a higher score indicates more symptoms of posttraumatic stress (47). The Swedish version has shown satisfactory psychometric properties (48).

*Depression symptoms* will be assessed with the Patient Health Questionnaire (PHQ-9), which consists of nine items describing symptoms of depression. Participants rate to what extent they experience these symptoms during the last two weeks on a 4-point scale (not at all – nearly every day, 0-3). The total score ranges from 0-27, and a higher score indicates greater symptom severity (49).

*Quality of life* will be measured with the Brunnsviken Brief Quality of Life Inventory, BBQ). It consists of 12 items covering six different life domains. Participants rate on a 5-point scale (strongly disagree – strongly agree, 0-4), to what extent they agree with the statement. Total score ranges from 0-96. The instrument is validated in a Swedish sample (50).

*Grief-specific avoidance* will be measured with the Depressive and Anxious Avoidance in Prolonged Grief Questionnaire (DAAPGQ). The questionnaire consists of nine items; five items measure depressive avoidance of activities and four items measures anxious avoidance of stimuli reminding of the loss. Participants rate on an 8-point scale (not at all true for me – completely true for me, 0-7) to what extent they experience the symptoms (51). Total score ranges from 0-72. The DAAPGQ was translated into Swedish by two grief researchers (JS & RE) at the Department of Neuroscience, Uppsala University in 2020 following European Organization for Research and Treatment of Cancer (EORTC) guidelines (52).

*Grief-specific rumination* will be measured with the Utrecht Grief Rumination Scale (UGRS). The instrument consists of 15 items measuring different aspects of grief rumination. The participants rate on a 5-point scale (never-very often, 1-5) how often they experienced certain thoughts over the last month (53). Total scores range from 15-75 and generate an overall grief rumination score. The instrument is validated in a Swedish sample with bereaved parents and indicates satisfactory psychometric properties (54).

*Negative grief cognitions* will be measured using a short version of Grief Cognitions Questionnaire (GCQ-18). It consists of 18 items, with statements regarding negative grief cognitions. Participants rate on a 6-point scale to what extent they agree with the statement (disagree strongly – agree strongly, 0-5). The total score ranges from 0-90 (55) and a higher score indicates stronger endorsement of negative cognitions (56). The GCQ was translated into Swedish by two grief researchers (JS & RE) at the Department of Neuroscience, Uppsala University in 2020 following EORTC guidelines (52).

#### Users’ experiences: perceived satisfaction, helpfulness and adverse events

A questionnaire with 25 items (9 of them with free-text response) regarding the user experience of the app will be used after participants have had access to the app for three months. The questionnaire includes questions regarding for example the perceived helpfulness and benefits of the app, which sections and tools the participants used most commonly and why, and if any parts of the app could be improved. Adverse events are assessed through the question “Did the use of the app have any negative consequences?” Participants who answered yes are asked to expand upon this.

### Data management

Study data is managed using REDCap electronic data capture tools (41) hosted at Uppsala University. All data will be de-identified with a code number. The key to the codes will be stored separately, and is only available to members from the research group.

### Statistical methods and analysis

T-tests and χ^2^ tests will be used to compare groups on all loss-related and sociodemographic variables. To examine intervention effects on the primary and secondary outcome variables, multiple regression models with the direct and interaction effect of condition × time with the primary endpoint at 3 months follow-up will be performed. Missing data at follow-up will be handled under the assumption of missing at random. Data of all randomized participants will be included in the analyses (i.e., intention-to-treat analysis). Cohen’s *d* and a 95% confidence interval will be calculated to assess the within and between-group effect sizes at post-intervention and follow-ups. Within group Cohen’s *d*’s, the standardized mean difference between the pre- and the post-assessments for each group, will be calculated (57). Between groups Cohen’s *d*’s will also be calculated by dividing the differences between change scores of both groups across time by the pooled standard deviation of both groups at baseline (58).

### Patient and public involvement

Two parents that lost their children to cancer were taking part in reading the content of the app and help out with translation of some questionnaires. The app will be pilot tested by 10-15 bereaved parents during April - May 2021. The parents will be asked to use *My Grief* for four weeks. After that, an in-depth assessment of the user experiences of the app will be conducted via telephone, by a member of the research team with questions, e.g., what it was like to use the app, how often they used it, which tools they used most commonly and why, and also whether they have suggestions to improve the app. The inputs and results of this testing may lead to changes in the intervention.

### Ethics and dissemination

All participants will fill out their consent to participate before enrolment, and they will be informed that they can decline their participation at any time without giving a reason. The pilot study (project no. 2020-01704) and the RCT (project no. 2021-00770) have received ethical approval from the Swedish Ethical Review Authority. Collected data will be handled confidentially, according to the European Union General Data Protection Regulation.

If the app is shown to be effective, the app will be made publicly accessible on app stores, so that it can benefit other bereaved people. Our findings will be presented to the Swedish Childhood Cancer foundation and other non-profit organizations for bereaved people. Colleagues will be informed about our findings during presentations at conferences and publications in scientific journals.

Taking part in this intervention could generate negative feelings and bring back negative memories, as it will elicit thoughts about the loss of the child. Any participant that reports experiencing harm or negative consequences as a result of their participation can, upon their request, be referred to an appropriate health care provider. However, we anticipate that participants can also experience positive outcomes, such as learning more about their own grief, increases in self-awareness, and improvements in mental health. Previous research shows that parents who lost a child to cancer valued participation in research as a positive experience despite extensive questionnaires of potentially sensitive character (59). Previous research also shows that participants who have experienced trauma show no negative effects of using the similar app PTSD Coach, and indicate that the app is both acceptable and feasible to use (42,44,45,60). Therefore, we believe that the benefits of participation in the study outweigh the risks.

## DISCUSSION

In this study protocol, we describe the development and planned evaluation of the self-management smartphone app *My Grief* for parents with prolonged grief symptoms after losing a child to cancer. This study will clarify whether the access of an app can have beneficial effects on bereaved parent’s mental health, quality of life and on cognitive-behavioral processes proposed to underlie the development of prolonged grief.

Offering a self-help app to bereaved parents in order to facilitate their grief process may have potential advantages, such as reducing barriers to seeking help, e.g., finding it too painful to speak about the loss, difficult to find appropriate help, or the negative effects of mental health stigma on help-seeking (13,61). The app is available free of charge to the users, and could be used in any geographic location and does not require visiting a health care organization. However, the *My Grief* app is not a guided treatment as it does not include any personal feedback from health care professionals or the research group, which could be perceived as a limitation. Studies on the PTSD Coach app have had dropout rates below 25% (36,45) and an ongoing RCT on the PTSD Coach Sweden, has a dropout rate of 11% (personal communication).

Generalizability of findings from this study may be limited for a number of reasons. Possibly, people who already received therapy or other social support prior to the loss may be more willing to participate in this study, because they may experience fewer barriers to seek help or talk about psychological problems (13,61). Similarly, people who are more willing to use technology, such as mobile apps or internet-treatments, would probably be more willing to participate in an app-based study (61–63). The generalizability of study outcomes may be affected in two ways by this selection bias. Tech-savvy users may appreciate the mode of delivery more which could have a positive impact on outcomes. Alternatively, those with more experience of technology could have higher standards and therefore higher demands of the app, which could affect their user experience and outcome in more negative ways. As seen in previous studies with the PTSD Coach app, some participants may experience difficulties with downloading or installing the app, or to remember to use it more than once during an ongoing intervention (64,65). In the *My Grief* app, a daily reminder can be set up to remind them to rate their grief in the grief chart self-assessment. The research group will also make a phone call to each user, about one week from registration, to check if everything is working. We expect that these measures will increase participant involvement in using the app. This study includes parents who have lost a child to cancer, hence the findings may not be generalizable to other causes, such as accidents and suicide, and more research is needed including parents who have lost a child due to other causes.

To conclude, this RCT will provide new insight to the effectiveness of a self-management app for prolonged grief in bereaved parents. If the app is found to be beneficial, it will provide a valuable addition to the bereavement support offered to bereaved parents in Sweden and may complement other psychological, medical and social interventions for prolonged grief. In addition, a next step could be to develop versions of the app for other groups of bereaved individuals and in other languages.

## Data Availability

Data will be available upon request to the principal investigator (JS) and unidentifiable data from this trial will be stored in data repositories at www.OSF.io.

## DECLARATIONS

### Competing interests

The authors declare that they have no competing interests.

### Funding

This research was funded by the Swedish Childhood Cancer Fund (PR2018-0047; TJ2018-0002). This funding source had no role in the design of this study and will not have any role during its execution, analyses, interpretation of the data, or decision to submit results.

### Author’s contribution

JS is principal investigator and grant holder. RE and JS are executive researchers and wrote the ethics proposals. RE, JS, ME, FA and PB developed the study design. RE, JS and ME developed the draft of the manuscript. FA and PB read, revised, and approved the draft of the manuscript. All authors approved the final manuscript.

## Acknowledgement

Thanks to the Veterans’ Affairs National Center for PTSD and Department of Defense’s DHA Connected Health for letting us base the *My Grief* app on the PTSD Coach source code. A special thanks to the two bereaved parents helping us with reading and commenting on the content in the app.

## Notes

### Competing Interest Statement

The authors have declared no competing interest.

### Clinical Trial

NCT04552717

### Author Declarations

Ethical approval has been received from the Swedish Ethical Review Authority (project no. 2021-00770).

